# In pursuit of the epileptogenic zone in focal epilepsy: A dynamical network biomarker approach

**DOI:** 10.1101/2022.02.15.22270978

**Authors:** Claudio Runfola, Hiba Sheheitli, Fabrice Bartolomei, Huifang Wang, Viktor Jirsa

## Abstract

The success of resective surgery for drug-resistant epilepsy patients hinges on the correct identification of the epileptogenic zone (EZ) consisting of the subnetwork of brain regions that underlies seizure genesis in focal epilepsy. The dynamic network biomarker (DNB) method is a dynamical systems-based network analysis approach for identifying subnetworks that are the first to exhibit the transition as a complex system undergoes a bifurcation. The approach was devised and validated in the context of complex disease onset where the dynamics is known to be nonlinear and high-dimensional. We here adapt and implement the DNB approach for the identification of the EZ from the analysis of SEEG data. The method is first successfully tested on simulated data generated with a large-scale brain network model of epilepsy using The Virtual Brain neuroinformatic platform and then applied to clinical SEEG data from focal epilepsy patients. The results are compared with those obtained by expert clinicians that designate the EZ using the Epileptogenicity Index (EI) method. High average precision values are obtained and posit the presented approach as a promising candidate tool for the pursuit of EZ in focal epilepsy.

**Author Summary:** We present a novel SEEG signal analysis tool for the identification of EZ regions in patients with drug-resistant focal epilepsy. The proposed method adapts and implements the dynamic network biomarker approach which builds on dynamical systems theory for complex networked systems. The method is first successfully tested on synthetic seizure data generated with The Virtual Brain modeling framework and then applied to retrospective patients’ clinical SEEG data. High precision values are obtained when the DNB subnetwork is compared with that designated as EZ by expert clinicians using empirical signal analysis measures and indicate that the DNB approach is a promising tool for the identification of EZ regions through SEEG signal analysis.

## Introduction

Surgical intervention is often a last resort lifesaving therapeutic option for patients with drug-resistant focal epilepsy. It consists in the anatomical localization and resection of the epileptogenic zone defined more than 50 years ago as the cortical zone at the origin of the seizures (Kahane et al., 2006; Talairach & Bancaud, 1966). The pre-surgical workup integrates non-invasive structural and metabolic imaging (MRI, PET scan) and electrophysiological data (video-EEG, MEG/EEG) to localize the epileptogenic regions and to search for a structural lesion. Very often invasive exploration is necessary and SEEG is the method of choice for this purpose (Isnard et al., 2018; Jehi et al., 2021). The focal (Rosenow & Lüders, 2001) or more distributed, networked nature of the epileptogenic zone (Bartolomei et al., 2017) has been much discussed in the past. The current extension of SEEG as the reference method (Jehi et al., 2021) brings a better understanding of electrophysiological phenomena. Ictal discharges are observed in many cases to occur simultaneously, or in rapid succession, in several distributed brain regions. In this context, the EZ concept has evolved to encompass what was now seen to be better characterized as a network of epileptogenic structures instead of a single focus, and in a sense, “… “focal” epilepsies are not in fact so focal but might instead involve distributed networks of varying spatiotemporal scales” (Bartolomei et al., 2017). Numerous quantitative approaches for network analysis were put forward in pursuit of the EZ (Bartolomei et al., 2017; Andrzejak et al., 2015). Examples include methods for analyzing functional connectivity via computation of linear or nonlinear measures of time-series correlations between brain regions or measures of causality for capturing directionality of interactions (Bartolomei et al., 2017). Graph theoretic approaches were also employed in attempts of identifying subnetworks comprising the EZ, often not without the challenge of finding clear interpretation of such measures in relevant neurophysiological terms (Bartolomei et al., 2017; Li et al., 2021). Network based statistics were also shown to provide promising features for training statistical learning algorithms that assign likelihood of individual regions belonging to the EZ (Li et al., 2018). While these empirical approaches provide useful and essential insight on seizure onset and propagation, further explanatory power remains to be sought from a connection to a formal mathematical theory of seizure dynamics. What we aim for in this work is to present a network-based method for EZ identification that builds on dynamical systems theory for the analysis of seizure dynamics. The dynamical systems framework (Jirsa et al., 2014) conceptualizes a seizure as a bifurcation in the neural dynamics through which the system undergoes a transition from a quiescent (healthy) state to an oscillatory (ictal) state. While seizure dynamics and propagation may be complex and high-dimensional, the seizure onset and offset are constrained by only a few degrees of freedom. Such is a consequence of the center manifold theory, which predicts that a small subset of system variables enslaves the system dynamics in the vicinity^1^ of the transition point (Guckenheimer and Holmes, 2006; Kelley, 1967; Carr, 1981). In the context of dynamics on networks, this predicts that when a bifurcation occurs, a subnetwork can be identified to be the first to move into the transition. This was exploited in (Chen et al., 2012) for the analysis of high-throughput omics data in the context of complex disease onset, such as data on the time evolution of genes or proteins expression whose dynamics are often modeled using nonlinear high-dimensional dynamical systems. A method was prescribed for identifying a dynamical network biomarker (DNB) as the group of nodes the dynamics of which will undergo a sudden strongly correlated change and that are the first to transit into the disease state. Given the genericity of the dynamical features which underlie bifurcations in networked nonlinear systems, such methods building on center manifold theory, or critical slowing down indicators, have found applications in various fields (Scheffer et al., 2009; Van De Leemput et al., 2014; Wichers et al., 2016; O’Regan et al., 2020; Brett et al., 2020; Kaur et al., 2020; Maturana et al., 2020). In our work here, we aim to repurpose and adapt the DNB method for the identification of the EZ subnetwork in focal epilepsy through the analysis of SEEG recordings.

The proposed method is first successfully tested with the use of synthetic data generated through The Virtual Brain Platform (TVB) (Sanz Leon et al., 2013). We then employ the developed algorithm for the analysis of SEEG data from patients with focal epilepsy and compare the results with those obtained empirically by expert clinicians. The resulting high precision values indicate that the DNB method is a promising new tool to be added to the armamentarium of epileptologists in the pursuit of the EZ in focal epilepsies.

## Results

The described algorithm was implemented for the identification of the EZ as a DNB subnetwork. It was first tested on a simulated dataset generated with the TVB modeling platform and then applied for the analysis of clinical SEEG time series.

### Simulated data

TVB was used to construct and simulate a large-scale brain network model of focal epilepsy. A subnetwork made of 5 ROIs was pre-set as the EZ by tuning excitability of 5 nodes to near threshold values. Another subset of nodes was tuned with subthreshold but high excitability to act as a propagation zone (PZ) subnetwork. The network was simulated and observed to undergo seizure-like discharges that originated in the EZ and propagated to the rest of the network. Figure 1A shows the simulation timeseries around seizure onset. The 5 nodes of the EZ are the first to spontaneously transit from baseline to ictal activity while the PZ regions follow, sequentially, relaying the seizure propagation through the network. A time interval for the analysis was chosen to encompass the passage into the ictal state, where the theory predicts the statistical properties of a DNB to arise. In figure 1B, the average SDs and pairwise absPCCs for the EZ group can be seen to undergo a sharp increase with seizure onset which leads to the amplification of the associated Composite Index *I*_*C*_, as predicted by the DNB method. In the same plots, statistical features of the PZ subnetwork also exhibit a substantial increase that is seen to lag behind that of the EZ group, in distinction with the rest of the nodes of the network that do not exhibit ictal activity. Note that if the EZ and PZ nodes were to be considered as one subnetwork, the composite index would exhibit multiple peaks that are attenuated in amplitude and not restricted to seizure onset, emphasizing the distinctiveness of the EZ signature when properly captured.

**Figure 1.**
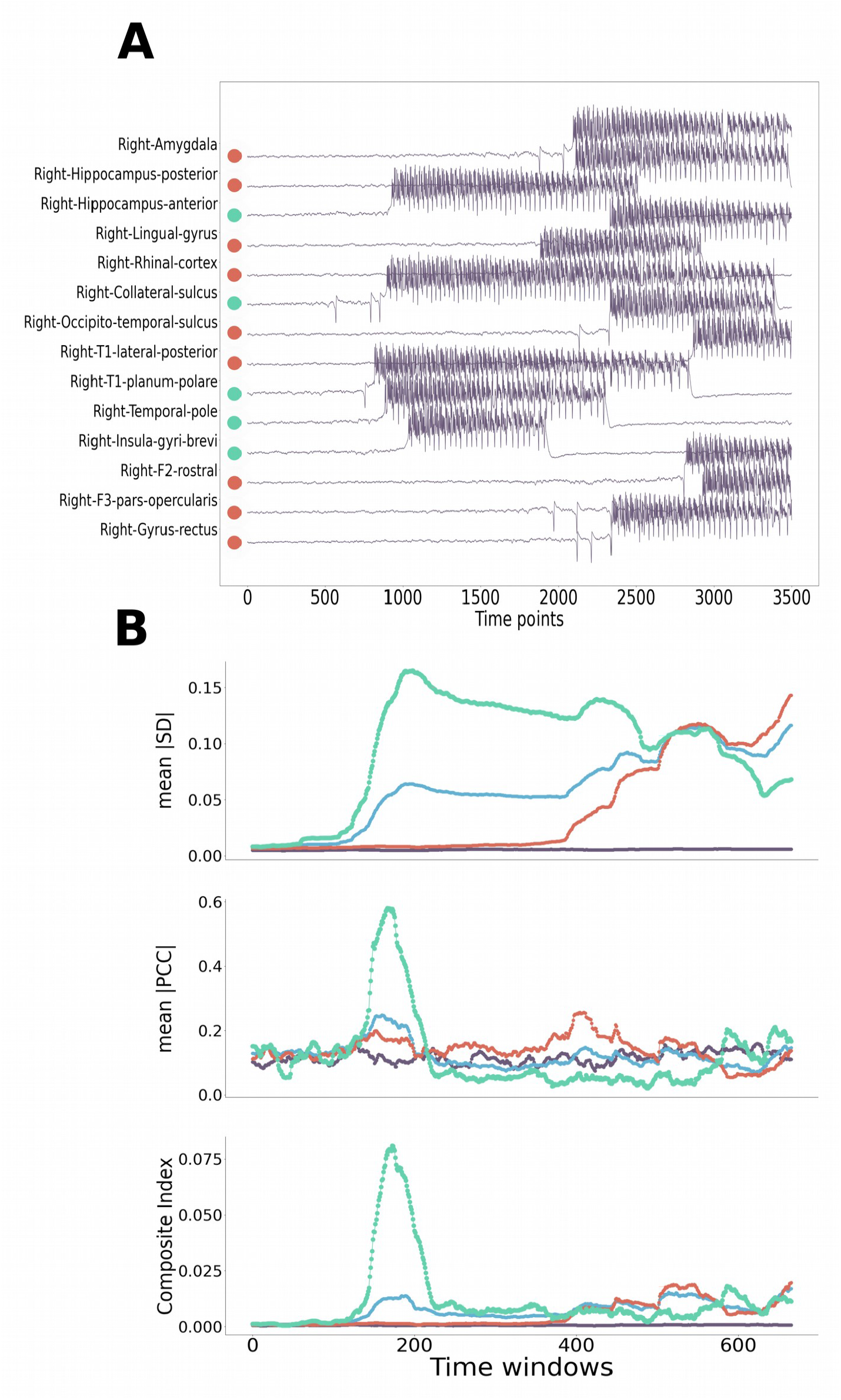
Applying the DNBs method to retrieve the EZ in simulated data. **A**) simulated time series of source activity for the EZ (green bullets) and PZ (red bullets) nodes. (**B**) the corresponding time series of the computed DNB statistical features for the EZ subnetwork (green), PZ subnetwork (red), EZ & PZ as one subnetwork (blue) and a sample subnetwork of NI nodes (not involved in the seizure) (purple).

Further analysis was applied to the computed DNB statistical features, as detailed in the Methods section, to attempt to identify the candidate subnetworks exhibiting increasing variance and growing correlations near the critical point. The contribution of individual nodes to the *I*_*C*_ of corresponding subnetworks was then screened to further prune the networks by excluding nodes of suboptimal contributions. The subnetwork with the highest resulting *I*_*C*_ was designated as the DNB subnetwork, that is, the EZ. The latter successfully matched the set of nodes that were tuned, by design, in the model to be of high excitability and capable of initiating ictal activity in the network.

### Clinical SEEG data

The developed algorithm was applied for the analysis of clinical SEEG recordings from 21 patients. The computations were performed on a relevant time interval of 10^3^-10^4^ data points centered around seizure onset times provided by expert clinicians. The neighborhood of the transition into ictal activity is then further narrowed down using statistical analysis (as detailed in the Materials and Methods section). The DNB statistical features were computed, and candidate DNB subnetworks were identified (as described in the Methods section). Nodes that were found to be included in the candidate subnetworks were further distilled by comparing overall contributions to the composite indices and then the resulting dominant group is finally designated as the DNB subnetwork, that is, the EZ.

To assess the performance of the method, we compare the EZ subnetwork compositions retrieved by the DNB method with those provided by expert clinicians. Precision was computed, as the ratio between true positives and the sum of true positives and false positives, in order to quantify the method’s ability to correctly identify nodes as belonging to the EZ. Figure 3A shows the distribution of precision values computed for individual patients; a patient’s dataset may present several seizures such that each data point here corresponds to an individual seizure for which the DNB method was applied. In figure 3C, precision values were averaged over seizure events and represented as one data point for each patient’s dataset. The results are then averaged over all patients and a mean precision value of 0.7 was obtained. When the EZ retrieved with the DNB method is compared against the union of the regions designated by the clinicians as EZ and PZ, the overall precision value increases to 0.9.

**Figure 2.**
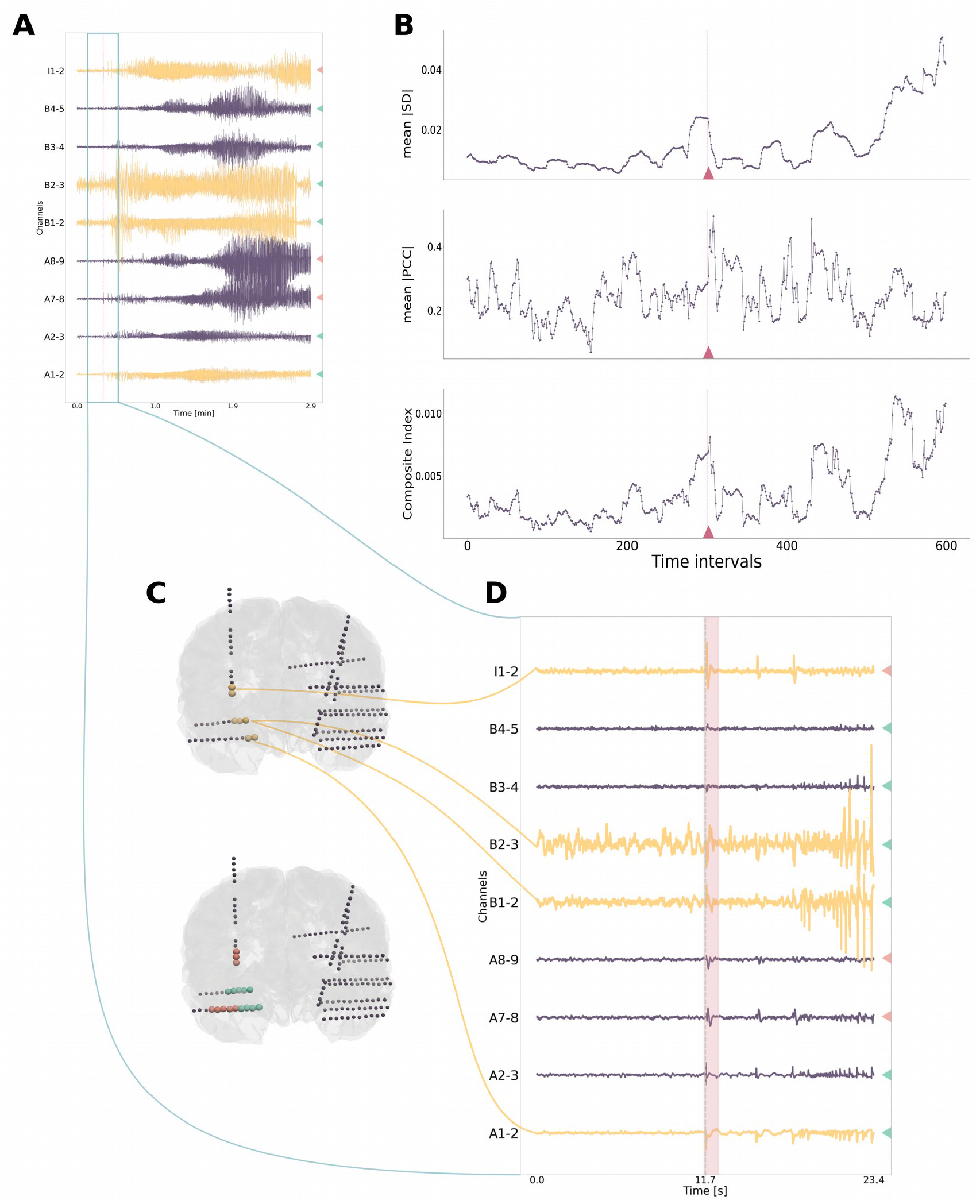
The DNB subnetwork analysis for a sample clinical SEEG seizure data. (**A**) Time series of electrode recordings with the largest SD in the neighborhood of the clinicians’ specified seizure onset time (light green window box); signals of the nodes of the subnetwork identified as DNB (in yellow); nodes designated by the clinicians as EZ and PZ, based on the Epileptogenicity Index (EI) method, are indicated by green and red triangles, respectively. (**B**) time series of the DNB statistical features for the identified DNB subnetwork in the analyzed window; the vertical red line marks the start of the interval of time where the DNB method indicates a seizure onset reflected by a sharp peak in the Composite Index. (**C**) schematic of the localization of the corresponding electrodes in the brain. (**D**) a zoom in on the neighborhood of the clinicians specified seizure onset time (same color scheme as in (A)); the pink window corresponds to the time interval indicated by the red triangle in (B).

**Figure 3.**
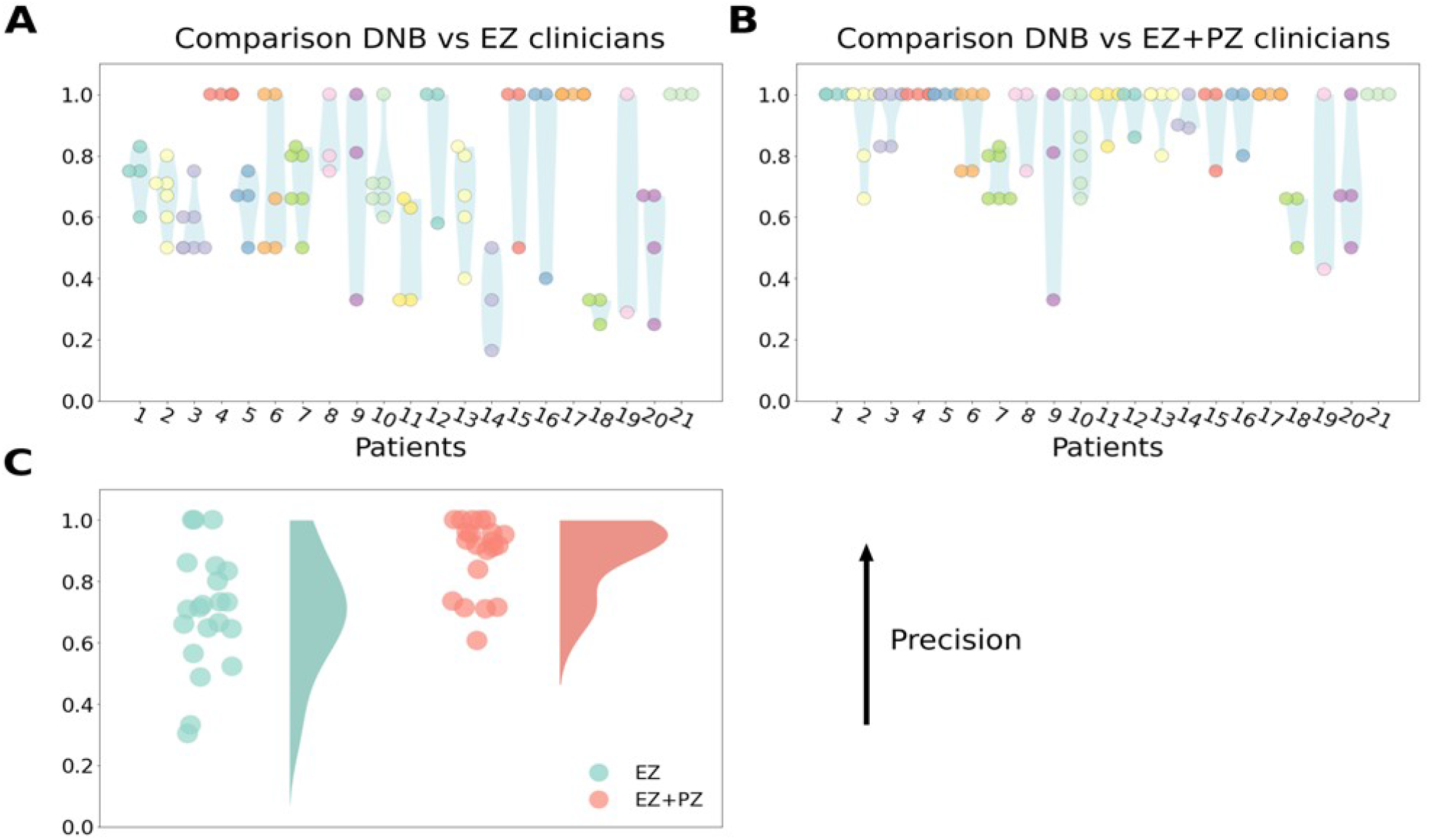
Precision values obtained by comparing results of DNB method to clinicians’ hypothesis (EI method). (A) precision value for each seizure of each patient (colored dots) as well as resulting distribution per patient (in light blue). (**B**) same as in (A) but comparing the DNB subnetwork to the union of EZ & PZ of clinician’s hypothesis. (**C**) average precision values per patient for the results shown in (A) & (B).

## Discussion

The prognosis after resective surgery in drug-resistant focal epilepsy is intricately dependent of the precise identification of an EZ subnetwork. By definition, the EZ is the ensemble of cortical regions that underlie the emergence of focal ictal activity and of which the removal, by surgical intervention, leads to the cessation of ictal activities, that is, to a patient’s freedom from seizures (Jobst et al., 2020). In 30-40% of all cases, seizures persist after surgery, and a major reason could be related to the fact that the EZ has not been correctly identified (Vaugier et al., 2018). This difficulty is partly caused by the distributed networked nature of the EZ. In the original work where the EI index was presented, when averaged over patients, the computed EI offered a clear distinction between the group of mesial structures (known to be involved in the EZ) and a second group of less epileptogenic lateral cortical structures, as values of EI obtained for the latter group were found to be significantly lower than those for the former group (Bartolomei et al., 2008). However, within the mesial structures themselves, a gradient of EI values was observed, supporting the fact that the EZ cannot be easily delineated to few unique structures but rather manifests as a complex network of distributed areas with variable epileptogenicity (Bartolomei et al., 2008). This further justifies the use of dynamical systems-based network analysis techniques in the analysis of seizure electrophysiological signals. The theoretical dynamical systems framework of seizure dynamics builds on a solid mathematical basis in bifurcation theory and has proven its power for the construction of personalized brain network models that are already potential tools for the guidance of epilepsy surgical interventions (Jirsa et al., 2017; Sanz Leon et al., 2013). Clinical approaches across many laboratories have validated the framework for seizure dynamics empirically and laid out a taxonomy of dynamotypes for a classification of seizures based on dynamics (Saggio et al., 2020). It was shown that the stereotypy of the main elements of seizure onset signals (fast oscillations and spikes) can be interpreted as an indicator of the existence of invariant dynamical properties shared by different seizures. Moreover, by characterizing the scaling behavior of frequency and amplitude during seizure onset and offset, specific bifurcation types can be identified as the underlying dynamical mechanisms through which these transitions into and out of seizure states occur.

Consistently, the DNB approach (Chen et al., 2012; Aihara et al., 2021; Zhan and Zhang, 2019; Liu et al., 2019; Vafaee, 2016) utilizes dynamical systems theory to provide a network analysis tool. The approach was devised for the identification of critical subnetworks that underlie the onset of complex diseases emerging from high-dimensional nonlinear dynamics in biological networks, as reflected in high throughput omics data. The relevance of the method for seizure dynamics arises from the commonality in dynamical features between large scale brain networks and the mentioned complex biological systems, which motivated our repurposing of the DNB method here for the identification of the EZ through the analysis of SEEG time series. The algorithm was first tested using the TVB modeling framework. This allowed for a validation against a ground truth as the mathematical model permits the pre-assignment of which nodes of the network are to be in the EZ subnetwork. Through the analysis of the simulated neural source data, the method succeeded in identifying the regions comprising the pre-assigned EZ subnetwork. As such, the evolution of the DNB statistical features for the simulated data was in line with what is conjectured based on the theoretical premises of bifurcation theory and the subsequent DNB approach. We then deployed the algorithm for the analysis of clinical SEEG data of drug-resistant focal epilepsy patients. The method showed good agreement with the EZ identification performed by expert clinicians as reflected in the resulting high average precision values. When the identified DNB subnetwork was compared against the union of the EZ & PZ from the EI method, the average precision increased substantially, indicating that the DNB method is differentiated from the EI method when it comes to distinguishing between EZ and PZ. Unlike in the case for simulated data, ground truth is unavailable here to perform further assessment. One approach to address this matter in future work can be the comparison of the results to post-operative MRI as a function of surgery success (Jirsa et al., 2017). In that context, a notable strength of the DNB method, in relation to previous relevant works for the EZ identification, is that the proposed algorithm has relatively very low computational cost and almost parameter free, except for the choice of sliding window. These two advantages well position it to be used for generating the prior for the more computationally intensive modeling-based methods such as the fully Bayesian inference methods behind the virtual epileptic patient (VEP) approach (Jirsa et al., 2017; Hashemi et al., 2021).

In conclusion, the results we present in this work provide a first proof of concept for the hypothesis that the EZ can be characterized as a subcritical network using dynamical network biomarkers (DNB). The performed computational analysis suggests that the DNB statistical features can be viewed as an indicator of seizure onset as their gradients exhibit sharp jumps with the approach of the system to a transition into the ictal state. The DNB method integrates concepts from dynamical systems theory with network analysis into a simple and computationally light method that can offer valuable insight for the identification of the EZ in focal epilepsy. Particularly, it emphasizes that the EZ can be viewed as a reflection of a center manifold arising around a bifurcation point in the system dynamics. The utility of the method in this context clearly illustrates that there is much to be gained in investigating epileptic phenomena with the lens of dynamical systems theory which serves as a crucial guiding principle for the theoretical analysis of large-scale brain dynamics.

## Materials and Methods

### The DNB composite index

The DNB method (Chen et al., 2012) integrates three main generic features of networked system dynamics that manifest in the vicinity of a critical transition (bifurcation point): 1) there exists a group of nodes, for which the average of the absolute values of their Pearson’s correlation coefficients (absPCCs) drastically increases. 2) the average standard deviations (SDs) of the activity of this group of nodes drastically increases. 3) the average of the absolute values of the Pearson’s correlation coefficients between nodes of this group and all other nodes drastically decreases. A composite index was then constructed to capture the relative extent of fulfillment of these criteria by groups of nodes of the network, and which will be maximal for the dominant group that is to be identified. For each candidate group of nodes, the composite index takes the form of a ratio in which the numerator is the product of the average SDs and the average of absPCCs for the group, and the denominator is the average absPCCs between the nodes of the group and all nodes not belonging to the group. In our study here, we use the same line of reasoning to construct a modified composite index (*I*_*C*_) that is equivalent to the numerator of that suggested in (Chen et al., 2012), that is, we focus on the first two generic features, such that *I*_*C*_=*SD*_*d*_. *absPCC*_*d*_, where *SD*_*d*_ is the average of the standard deviation of the elements in the dominant group and *absPCC*_*d*_ is the average of the absolute values of the Pearson’s correlation coefficients of the paired elements in the dominant group. The modification is reasoned to enhance discrimination between nodes of the EZ and those of what is often referred to as the propagation zone (PZ), that is nodes through which the seizure originating in the EZ propagates onto the rest of the network, and are expected to be distinct from but have high absPCC values with those of the EZ.

### The brain network model

The Virtual Brain platform (Sanz Leon et al., 2013) was used to construct a large-scale brain network model that is capable of generating timeseries datasets with the basic features of epileptic brain network activity as observed in SEEG. Structurally, the simulated brain is made of several brain areas (nodes) according to the parcellation of the Virtual Epileptic Patient atlas (Wang et al., 2021) which includes 162 regions of interest (ROI) that represent 73 cortical and 8 subcortical structures per hemisphere. Connectivity between the nodes of the network was prescribed by a sample structural connectome obtained from tractography analysis of diffusion weighted imaging (Wang et al., 2021). The activity of each node was modeled using the *Epileptor* (Jirsa et al., 2014) model which is a phenomenological model devised to capture the canonical dynamical features of ictal dynamics. The model allows for the tuning of the epileptogenicity of each node via an excitability parameter, *x*_*0*_, such that a region will exhibit ictal discharges for a critical threshold value, *x*_*0C*_. Accordingly, we designate a subset of the nodes to be in the EZ by setting *x*_*0*_ *=-1*.*325*, which is just below the threshold, and another subset is tuned to act as a propagation zone (PZ) with *x*_*0*_ *=-1*.*775*. In particular, in the model, the EZ is chosen to include five ROI matching those obtained by clinicians for a sample dataset, while the PZ is chosen to be composed of ten ROI that are randomly selected nodes which are highly connected to the EZ. Additive Gaussian noise is included in the model to allow for small random fluctuations in the dynamics. SEEG-like signals were obtained as linear projection of the Epileptor variables (‘x_2_ - x_1_’) from a 10000 timesteps time series simulation. The signal is then normalized by dividing by the difference between its maximum and its minimum.

### Signal analysis

The analysis of the obtained signals was done using Python programming language. To compute the various statistical quantities required for the implementation of the DNB approach, such as correlations and variances, the time series were analyzed using a sliding window approach. Starting from the pre-ictal state, intervals of around 1000 timesteps were analyzed with a sliding window of 330 timesteps and a sliding shift of 4 timesteps. For each window, a Functional Connectivity (FC) matrix was constructed whose elements are the absolute values of the pairwise Pearson’s correlation coefficients between ROIs. Standard deviation (SD) vectors were computed for each ROI in each window. The resulting tensors are of dimensions N_regions_ x N_regions_ x N_windows_ and N_regions_ x N_windows_, for FC and SD respectively; N_regions_ is the total number of ROIs in the parcellation and N_windows_ is the total number of sliding windows in the analyzed time interval.

### Retrieving the EZ as the DNB subnetwork

The DNB composite index computations were done for the time interval encompassing the vicinity of the seizure onset. For the clinical SEEG data analysis, the seizure onset zone was identified by expert clinicians through visual analysis and epileptogenicity index computation (Bartolomei et al., 2008). For the simulated data, the time interval to be analyzed was that in which the SDs and absPCCs of the most active nodes are maximum in concurrence with a maximum reduction of the variance explained by the first few principal components of the time series. Intuitively, this corresponds to looking for the concurrence of the conditions for a DNB subnetwork with a drastic decrease in the effective dimensionality of the system dynamics, as a signal for near proximity to a transition point (see Supplementary Materials for details). For computational efficiency, we focus the analysis on the ROIs that are found to be exhibiting dynamics with variance values within the top percentile for more than two-thirds of the time. Pairs of ROIs with increasing correlation, reflected by positive average successive increments in absPCC values, are grouped into corresponding subnetworks to be further screened by the DNB criteria. For each of these candidate subnetworks, the maximum value of the modified DNB composite index (*I*_*C*_) was computed. A matrix *I*_*ij*_ is then constructed whose elements are the contribution of node *i* to the *I*_*C*_ of subnetwork *j* (*I*_*ij*_*=0*, if *i* is not in *j*). Each candidate subnetwork is then pruned by excluding the nodes whose sum of the absPCCs with the nodes of the group is less than 50% of that of the nodes of the group among each other. The candidate subnetwork with the highest resulting *I*_*C*_ is then taken to be the DNB subnetwork, that is, the EZ for the analyzed seizure.

### Clinical SEEG data

The described selection algorithm was applied to real ictal SEEG data from 21 retrospective patients with drug-resistant focal epilepsy, whose pre-operative evaluation was done at La Timone hospital, Marseille (Wang et al., 2021). Signals were acquired by SEEG invasive recordings from multiple depth electrodes, with 10-18 contacts of 2 mm length and 1.5 or 5 mm spacing, collected on a 256 channel Deltamed/Natus system, without digital filters. During the acquisition, a high-pass filter (0.16 Hz of cut-off frequency at – 3dB) and an anti-aliasing low-pass filter (a third of the sampling frequency as cut-off) were applied. The position of the electrodes was obtained by a cranial CT scan after the implantation. CT-scan/MRI data co-registration was performed, to check the anatomical location of each contact along the electrode’s trajectory, using the open-source software GARDEL ((Medina Villalon et al., 2018). For each patient, clinicians associated to each bipolar channel a ROI tagged as EZ, PZ or non-identified (NI), based on the Epileptogenicity Index method (Bartolomei et al., 2008), full details on the data acquisition and EZ designation by clinicians are provided in (Bartolomei et al., 2008; Contento et al., 2021). Time intervals of the order of one to 20 seconds around the clinicians specified seizure onset time were analyzed. A one second time window (with a sliding shift of 10 data points) was used for the statistical computations. The analysis is first narrowed down to that of nodes having the highest variance for at least 60% of total time and sharp peaks in their mean composite index are used as complementary indicators of transition points. The developed algorithm is then applied to identify candidate DNB subnetworks as done for the simulated data. The distinction between EZ and PZ is often more challenging in clinical data than in simulated data as the signals in the former are more complex. For further screening of the candidate subnetworks, we construct a matrix whose elements, *Wij*, are the contribution of node *i* to the composite index of candidate subnetwork *j*. The nodes with a cumulative contribution that is higher than the mean value of *Wij* are taken to constitute the dominant DNB subnetwork corresponding to the EZ.

### The Epileptogenicity Index

The EZ subnetwork for each analyzed seizure, as identified using the proposed DNB approach, is compared with those obtained by expert clinicians using the measure of the “Epileptogenicity Index” (EI). Generally, for the identification of regions of the EZ, clinicians rely on two key features of ictal signals. The first is the propensity of a given region to generate high-frequency oscillations (rapid discharges) and the second is the relative timing of a given region’s engagement in the seizure activity, that is, the sooner (closer to onset) a given region exhibits rapid discharges, the more likely it is to be designated as part of the EZ. The EI is a quantitative metric that was devised to capture those two key features by combining a quantification of the ictal spectral and temporal properties of recorded brain structures (Bartolomei et al., 2008). This measure of the epileptogenicity of brain regions was first developed and validated in the context of mesial temporal lobe epilepsies, which is known to involve a “multistructural epileptogenic zone”. We refer the reader to the original work (Bartolomei et al., 2008) for full details on the computation of EI.

## Data Availability

Patients raw datasets cannot be made publicly available due to the data protection concerns regarding potentially identifying and sensitive patient information. Interested researchers may access the datasets by contacting the corresponding authors

## Acknowledgements

Support was provided by the French National Research Agency ANR as part of the second “Investissements d’Avenir“ program, ANR-17-RHUS-0004, EPINOV (https://anr.fr/) and the European Union’s Horizon 2020 Framework Program for Research and Innovation under the Specific Grant Agreement No. 785907 (https://ec.europa.eu/programmes/horizon2020/en).

## List of technical terms

- Seizure: a transient occurrence of signs and/or symptoms due to abnormal excessive or synchronous neuronal activity in the brain
- Bifurcation: a sudden qualitative change in the system dynamics as a critical value of a parameter is crossed.
- Critical slowing down: the propensity of a system to exhibit an increasingly slow recovery from perturbations as it approaches a bifurcation point.
- Pearson’s correlation coefficients: the ratio of the covariance of two variables to the product of their standard deviations; provides a measure of linear correlation between two datasets.
- Center manifold: intuitively can be understood as the subspace spanned by the directions along with trajectories are neither attracted nor repelled in the neighborhood of an equilibrium point of a system; a subspace spanned by the eigenvectors of the Jacobian, at an equilibrium point, corresponding to the eigenvalues with zero real part.

## Footnotes

1 Vicinity here can be intuitively understood as a brief interval that extends from just before to just after the transition

