## Supplementary Material for "In pursuit of the epileptogenic zone in focal epilepsy: A dynamical network biomarker approach"

**Procedure to narrow down the time window to a neighborhood of the transition point.**

For practical purposes, it was necessary to narrow down the analysis to an interval of time centered around what we refer to as the working point, which can be thought of intuitively as the time window in which the DNB statistical features are most discernible. Regions whose standard deviation is the highest for about the 20% of the whole time series are observed. The time series of the average SD and absolute value of PCCs of these nodes are monitored for sharp peaks to be detected. Principal Component Analysis (PCA) is performed for each time window to reduce the dimensionality reduce of the signal. Peaks in the time evolution of the proportion of variance explained by the first two components (PVE2) are detected. In addition, the dynamical Functional Connectivity (dFC) (Hansen et al., 2015) is computed to detect sharp peaks in the PCCs. The working point interval is then taken to be that in which all the SD, PCC, dFC and PVE2 densities are maximally overlapping.

---

* These authors contributed equally to this work.
